# Inequalities in ownership and availability of home-based vaccination records in 82 low- and middle-income countries

**DOI:** 10.1101/2024.01.16.24301369

**Authors:** Bianca de Oliveira Cata-Preta, Andrea Wendt, Thiago Melo Santos, Luisa Arroyave, Tewodaj Mengistu, Daniel R Hogan, Aluisio JD Barros, Cesar Gomes Victora, M. Carolina Danovaro-Holliday

## Abstract

**Introduction:** Home-based records (HBR) are widely used for recording health information including child immunizations. We studied levels and inequalities in HBR ownership in low- and middle-income countries (LMICs) using data from national surveys conducted since 2010.

**Methods:** 465,060 children aged 6-35 months from 82 LMICs were classified into four categories: HBR seen by the interviewer; never had an HBR; had an HBR that was lost; and reportedly have an HBR that was not seen by the interviewer. Inequalities according to age, sex, household wealth, maternal education, antenatal care and institutional delivery were studied, as were associations between HBR ownership and vaccine coverage. Pooled analyses were carried out using country weights based on child populations.

**Results:** An HBR was seen for 67.8% of the children, 9.2% no longer had an HBR, 12.8% reportedly had an HBR that was not seen and 10.2% had never received one. The lowest percentages of HBRs seen were in Kiribati (22.1%), the Democratic Republic of Congo (24.5%), Central African Republic (24.7%), Chad (27.9%), and Mauritania (35.5%). The proportions of HBRs seen declined with age and were inversely associated with household wealth and maternal schooling. Antenatal care and institutional delivery were positively associated with ownership. There were no differences between boys and girls. When an HBR was seen, higher immunization coverage and lower vaccine coverage rates were observed, but the direction of this association remains unclear.

**Interpretation:** HBR coverage levels were remarkably low in many LMICs, particularly among children from the poorest families and those whose mothers had low schooling. Contact with antenatal and delivery care was associated with higher HBR coverage. Interventions are urgently needed to ensure that all children are issued HBRs, and to promote proper storage of such cards by families.

## Introduction

Home-based records (HBR) are personal health documents used to record the history of health services received by an individual.(1) The most frequently used HBRs are child vaccination cards issued by health services at the time of the first vaccination - often in the hospital or facility where the child was born – and usually kept in paper format by the child’s caregivers. The use of HBRs that include vaccination status is widespread – being implemented in at least 163 countries as of 2018(1) - but HBRs vary greatly in design and regarding the information documented, which includes growth curves and clinic visits in several countries. This article refers to HBRs as documents that record vaccines received by children, with or without additional health data.

HBRs are important tools for several reasons.(1) HBRs serve as a documentation tool that allows parents and health services to track the immunization status of a child, and to support compliance with vaccination schedules. They will facilitate continuity of care by providing a reliable source of information for healthcare providers, thus allowing providers to quickly determine which vaccines a child has already received and when they are due for the next vaccine. For families with limited access to services, such as those living in remote areas, HBRs may help schedule the travel necessary to obtain vaccines. HBRs are also important for monitoring immunization coverage through household surveys.

Although evidence of the effectiveness of HBRs in increasing vaccination coverage is scant,(2) there are studies suggesting improved timeliness and reduced missed opportunities for vaccination.(3, 4) In fact, a World Health Organization (WHO) committee produced a positive appraisal of HBRs, noting that these contribute to the right to access to information and support global efforts for people-centred care.(1) Furthermore, in early 2023, WHO published updated guidance to strengthen the use of HBRs as a tool improve care seeking behaviours, maternal and child home care practices, infant and child feeding, men’s involvement and support in the household, and communication between health workers and women, parents and caregivers.(5) HBRs are also important for vaccination status ascertainment in surveys - as in their absence the information may be affected by recall bias. This has an important impact on estimates of immunization coverage, as the low availability of HBRs has been highlighted as a key challenge for vaccination coverage surveys.(6) (7, 8)

A 2015 analysis found that the proportions of children for whom HBRs were available for inspection was under 50% in 24 of 67 of low and middle-income countries (LMICs) studied.(9) Low HBR coverage in many LMICs highlights the need for investigating which children are most likely to lack such records.

Substantial inequalities in child immunization coverage have been widely documented in LMICs.(5, 10–13) Well-established determinants of coverage include maternal education, household wealth and geographical location, whereas inequalities according to the child’s sex are not so evident.(10–12, 14) Ethnic group affiliation(15, 16) and religion(16) are also determinants of vaccine coverage in some, but not all countries. To our knowledge, the association between ownership of HBRs and immunization outcomes has not been systematically studied using survey data.

In contrast to the ample literature on immunization coverage inequalities, we were unable to identify any single-country or multicountry analyses on disparities in HBR ownership. The factors affecting HBR ownership in LMICs are likely the same as those affecting immunization coverage. In addition to the above-described characteristics of the household and the child, coverage is driven by health systems-related factors that include HBR and vaccine supply, distribution logistics and facility readiness, as well as staff availability and training.(5, 17, 18) Uptake is also influenced by factors such as vaccine hesitancy(19, 20) and political instability.(21)

Analyses of HBR coverage have so far focused on the proportion of children who, during a household survey, have an HBR in the home that was shown to the interviewer. However, there are other possibilities. Children for whom an HBR was not available for examination may have never had such a card; have once had an HBR but no longer have one; or reportedly still have an HBR, but the interviewer could not see it. This distinction is important because such results may provide insights on how to expand the use of HBRs and improve measurement of vaccine coverage. Lastly, there are also countries where vaccination records are kept in health facilities and are not available for inspection during a household survey.

In this article, we use nationally representative surveys with information on vaccination carried out between 2010 and 2021 to describe the distribution of children according to the four categories of HBR ownership listed above. We also show how these proportions vary by country, child’s age and sex, and selected sociodemographic and healthcare characteristics. In addition, we explore whether HBR status is associated with survey-based measures of vaccine coverage and drop-out between DPT1 and DPT3. Our aim is to better understand the spectrum of HBR status to inform how to improve their ownership, which could translate in increased demand for immunization, better timeliness of vaccination, and - at the health systems level - improved monitoring and measurement of vaccination status.

## Materials and Methods

The International Centre for Equity in Health database includes all publicly available datasets from nationally representative Demographic and Health Surveys (DHS) and Multiple Indicator Cluster Surveys (MICS).(22, 23) Over 450 surveys from 125 countries were screened to select the most recent survey from each country carried out since 2010. Of these, 82 with information on ownership of HBRs and on immunization history (47 DHS and 35 MICS) were identified and are part of this study. Eleven other countries where vaccine records were only kept in health facilities were excluded (Ethiopia, Kazakhstan, Kyrgyzstan, Moldova, Mongolia, North Macedonia, Serbia, Tonga, Turkmenistan, Ukraine and Vietnam). Further information on the surveys is available elsewhere: DHS (https://dhsprogram.com/what-we-do/survey-Types/dHs.cfm) and MICS (http://mics.unicef.org/). DHS and MICS are highly comparable in terms of sampling and indicators.(24, 25)

Our study sample included children 6-35 months as most surveys did not collect immunization information on older children. Those aged under six months were excluded because they may not yet have been issued an HBR. Four categories of HBR status were analysed: 1) HBR seen by the survey interviewer (“HBR seen”); 2) HBR reportedly existed but was not produced for inspection (“has HBR, but not seen”, also referred to as “misplaced HBR”); 3) HBR available at some point but had been lost (“no longer has an HBR”); and 4) HBR never issued to the child (“never had an HBR”).

We investigated associations of HBR ownership with the following variables:

- Age (6-11, 12-23 and 24-35 months) and sex of the child.
- Household wealth quintile, derived from household asset indices provided in the DHS and MICS datasets. These were obtained using principal component analyses of household assets and characteristics of the building, presence of electricity, water supply and sanitary facilities, among other variables associated with wealth. Because relevant assets may vary in urban and rural households, principal component analyses are initially carried out separately for each area and later combined into a single score using a scaling procedure to make area scores comparable.(26)
- Maternal schooling in three categories, as reported by the survey respondent: none (no formal schooling); primary (any primary schooling, including completed primary education); secondary or higher (any secondary education, including completed secondary schooling and partial or full higher education).
- Area of residence coded as urban or rural according to country-specific delimitations.
- Four or more antenatal care visits to any provider during the pregnancy. For both DHS and MICS, this information is available for the lastborn child only, who account for 76.7% of our study sample.
- Institutional delivery, defined as delivery in a health facility of any level. DHS collects this information only for lastborn children, whereas MICS provides information on all under-fives. Data are available for 87.8% of the study sample.

Analyses of the association between HBR ownership and vaccine coverage were done with BCG (one dose), poliomyelitis (three doses, either oral or injectable), diphtheria-pertussis-tetanus (DPT) (one and three doses of any DPT-containing vaccine), and measles (one dose of any measles-containing vaccine or MCV). Full immunization coverage was defined as receiving at least one dose of each BCG and MCV, and three doses of each DPT and polio. The DPT dropout ratio was defined as the percentage of children who failed to receive a third dose among those who received the first dose. The same approach was used to calculate the DPT1 to MCV dropout ratio, even though having received DPT1 is not a prerequisite for receiving MCV.

The association analyses with vaccine coverage were restricted to children 12-23 months; in countries where MCV is recommended at ages 15 or 18 months, the analyses included children aged 15-26 or 18-29 months, respectively. The information on vaccines was collected from HBRs or, if not available for inspection, from the caregiver’s recall. Children without any information on vaccination status were considered as unvaccinated.

The units of analyses were all children aged 6-35 months from the 82 countries with data. Analyses of each survey dataset accounted for the multi-stage survey design and sampling weights. The pooled analyses included multinomial regression with HBR as a four-category outcome including fixed effects for country, and were weighted by the populations of children aged 12-23 months in each country as of 2017 (median year of all surveys), according to the World Bank.(27)

Analyses were stratified by World Health Organization regions: African Region (AFR), Region of the Americas (AMR), South-East Asian Region (SEAR), European Region (EUR), Eastern Mediterranean Region (EMR) and Western Pacific Region (WPR) and by country income (low, lower-middle or upper-middle income) using the most recent World Bank classification.

The analyses were carried out with Stata (StataCorp. 2019. Stata Statistical Software: Release 17. College Station, TX: StataCorp LLC) and R (R Core Team, 2020, version 4.1.0. R Foundation for Statistical Computing, Vienna, Austria).

Ethical clearance was the responsibility of the institutions that administered the surveys and all analyses relied on anonymized databases.

## Results

We studied 465,060 children aged 6-35 months; 20% were aged between 6 and 11 months (93,411 children), 40.0% (185,351) between 12 and 23 months and 40.1% (186,298) between 24 and 35 months. We included 82 countries representing 82% of all low income, 62% of all lower-middle income, and 45% of all upper-middle income countries as of 2017. Supplementary Table 1 lists all study surveys. Information on HBR, sex, age, wealth quintiles and area of residence was available for all children. Maternal education, antenatal care and institutional delivery were not available for 36, 108,481 and 56,767 children. Vaccine coverage analyses were restricted to 185,729 children aged 12-23 months.

Across world regions, EUR countries presented the highest median value for children whose HBR was seen at the survey interview (89.6%), while the WPR countries presented the lowest median (59.0%). The lowest values were in Kiribati (22.1%) and in four AFR countries

(Democratic Republic of Congo (24.5%), Central African Republic (24.7%), Chad (27.9%), and Mauritania (35.5%)).

EUR presented the lowest national median values in the “has HBR, but not seen” group (6.2%), no longer having an HBR (0.9%), and never having had an HBR (1.0%). Median values for no longer having an HBR varied from 0.9% in EUR to 9.6% in WPR, whereas the medians for never having an HBR ranged from 1.0% and 1.1% in EUR and AMR, respectively, to 9.7% in WPR. Table 1 shows HBR ownership status according to the six WHO regions.

**Table 1.**
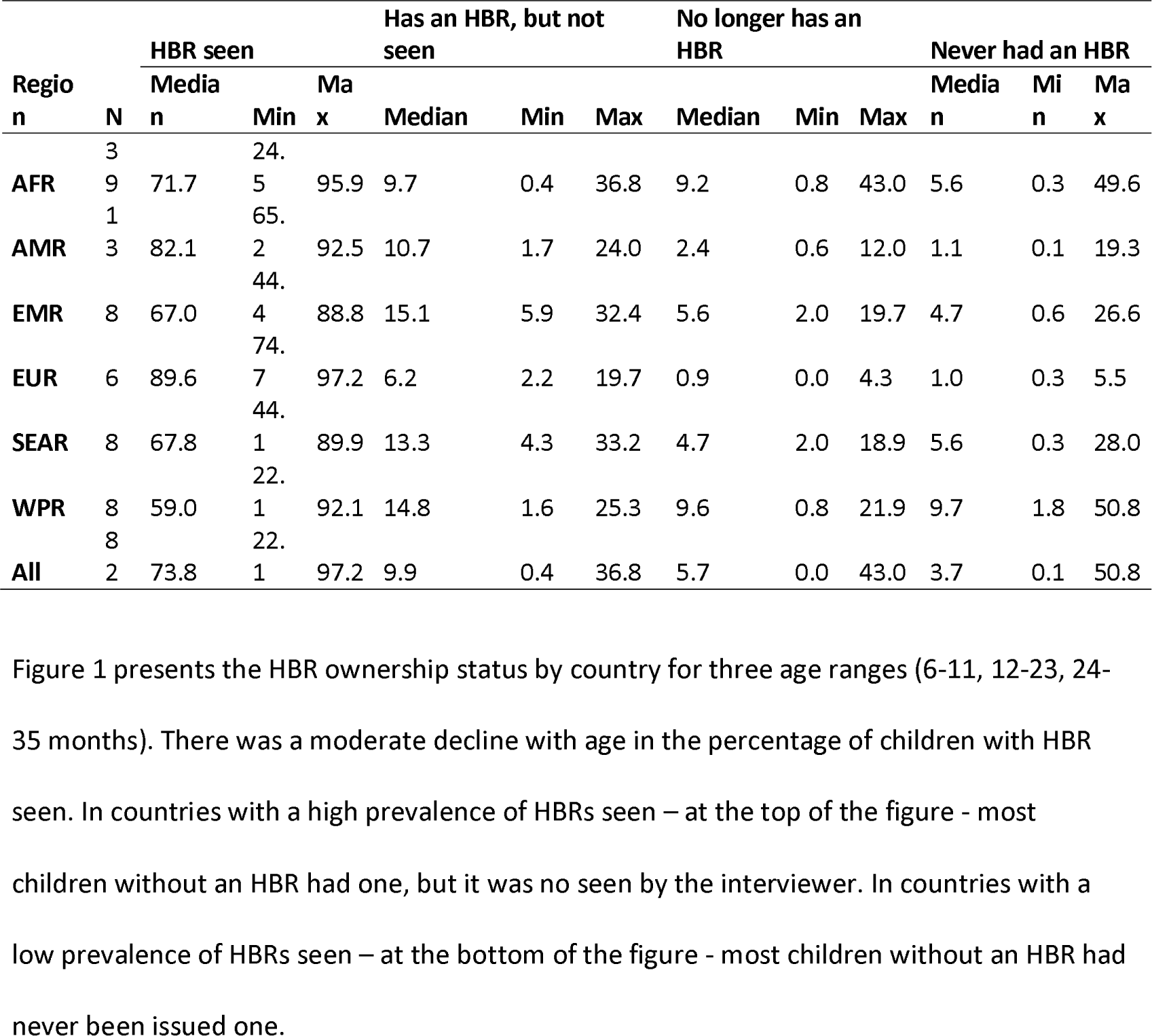
HBR ownership for children aged 6-35 months according to WHO regions.

Figure 1 presents the HBR ownership status by country for three age ranges (6-11, 12-23, 24-35 months). There was a moderate decline with age in the percentage of children with HBR seen. In countries with a high prevalence of HBRs seen – at the top of the figure - most children without an HBR had one, but it was no seen by the interviewer. In countries with a low prevalence of HBRs seen – at the bottom of the figure - most children without an HBR had never been issued one.

**Figure 1.**
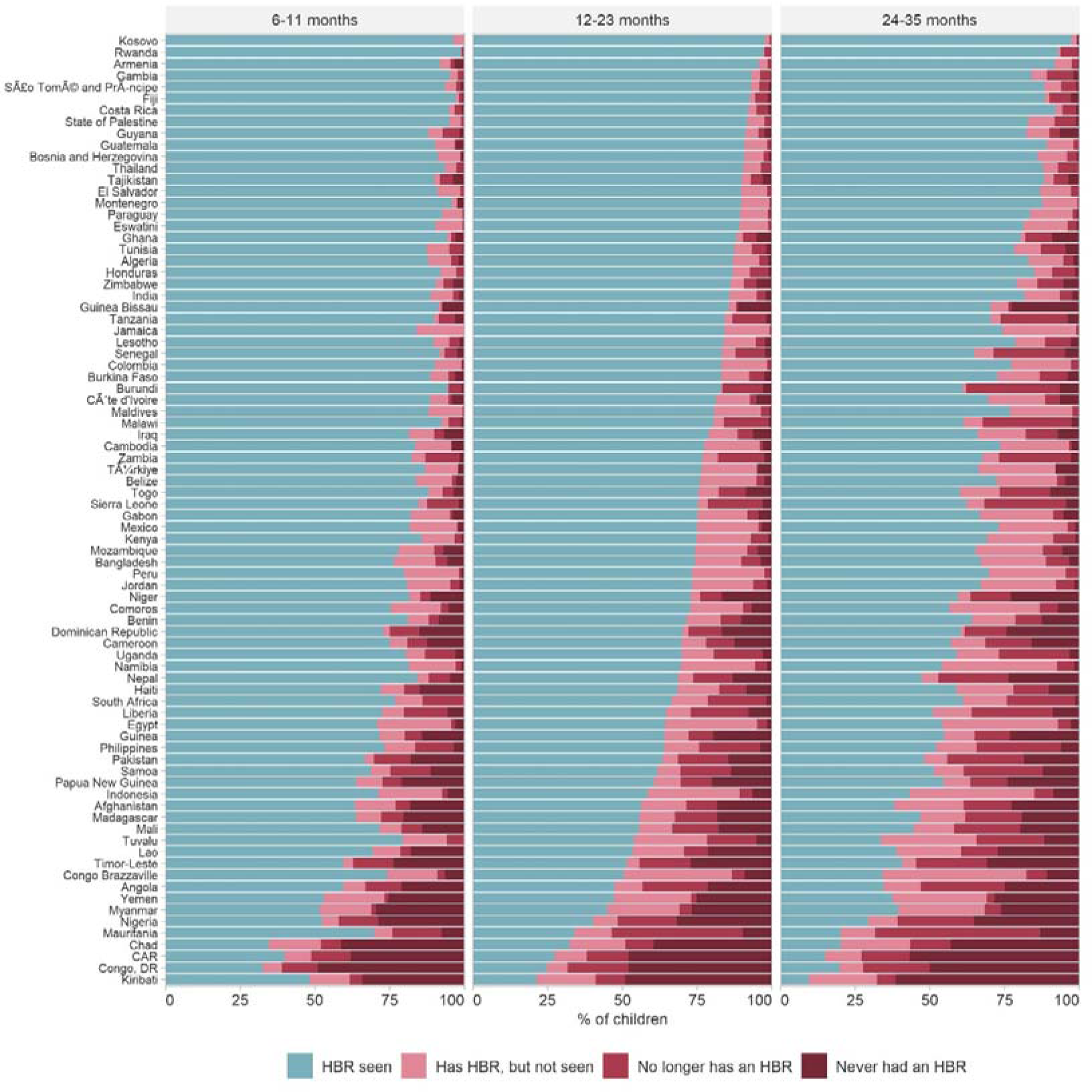
Percentage of children according to HBR status by country. Countries ordered according to the % of children aged 12-23 months with HBR seen

Figure 2 summarizes the results from the pooled analyses of the 82 countries across inequality dimensions (see Supplementary Table 2 for more detailed results, including 95% confidence intervals). After weighting by national child population, 67.8% of the children aged 6-35 months had an HBR seen by the interviewer, 9.2% no longer had an HBR, 12.8% reportedly had an HBR that was not seen and 10.2% had never received one.

**Figure 2.**
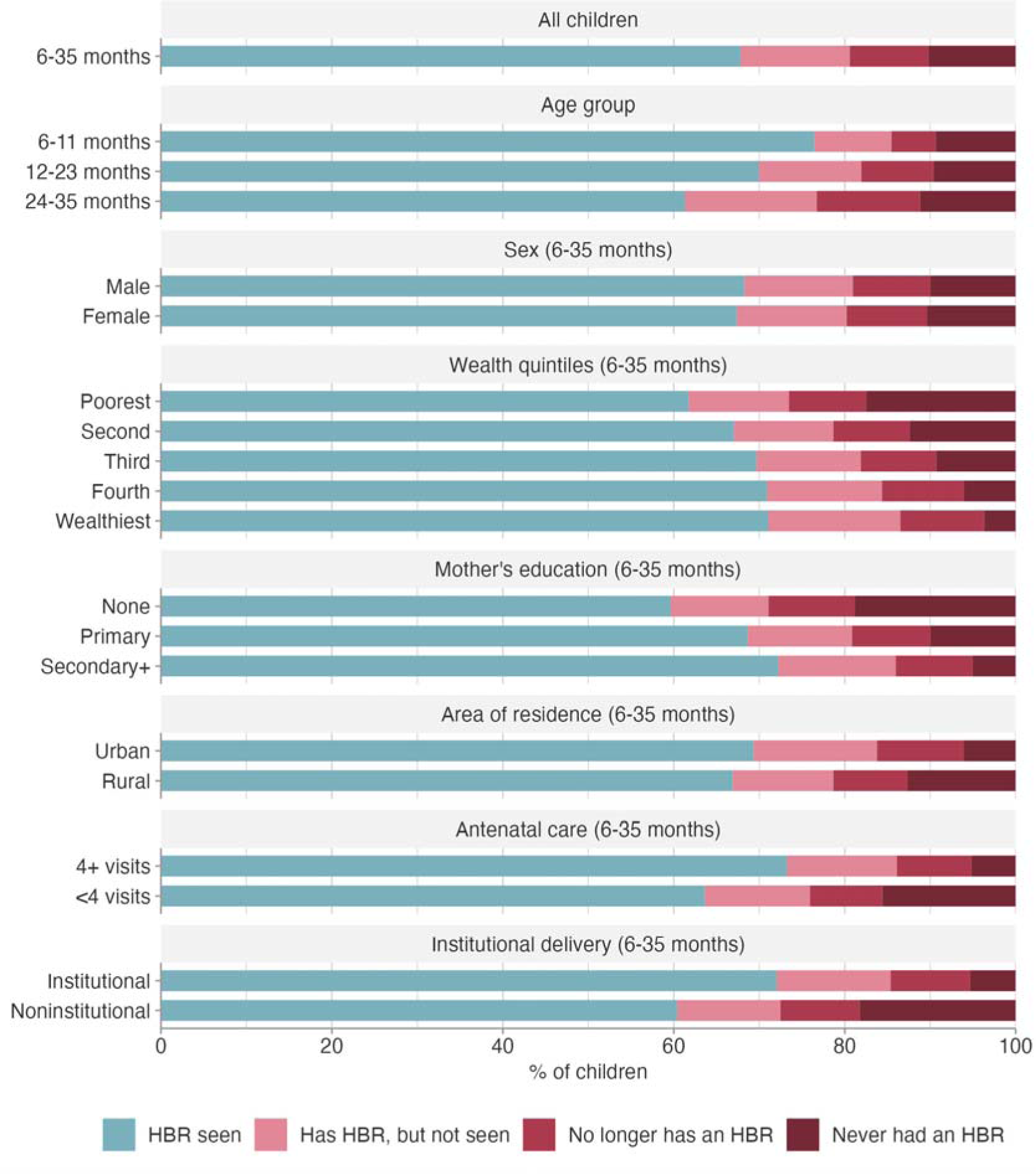
HBR ownership by characteristics of the child and household.

Figure 2 also shows inequalities in HBR ownership. The prevalence of HBRs seen dropped rapidly with the age of the child, with increasing proportions of children who no longer had an HBR or children who had an HBR that was not seen. These cross-sectional results suggest that 0.7% of HBRs are lost or misplaced with each increasing month of age (P<0.0001), but the proportion of children who never had an HBR did not vary with age (P=0.191). There were virtually no differences between boys and girls.

In terms of household wealth, the percentages of children with HBRs seen increased monotonically from 62.4% in the poorest quintile to 70.1% in the wealthiest quintile. Conversely, 3.7% of the wealthiest children never had an HBR compared to 17.1% among the poorest. This gradient is even more evident in terms of maternal education. While HBRs were seen for 73.4% of children born to women with at least secondary education, this proportion was down to 55.7% when the mother had no formal schooling. Among the latter group, 23.0% never had an HBR, compared to 4.3% among the former.

The percentages of children with HBRs seen by the interviewer were very similar for rural and urban residents, but the percentage of rural children who never had an HBR (12.4%) was twice as high as that of urban children (6.3%), and conversely more urban than rural children reportedly had an HBR that was not seen.

We also investigated the overlap between HBR ownership and healthcare utilization. Higher proportions of children with HBRs seen were observed when their mothers attended antenatal care four or more times than those with fewer or no visits (73.2% and 63.1%, respectively), a group in which about one-fourth of the children never had or no longer have an HBR. Similarly, HBRs seen were more common among children delivered in a health facility than those born at home (74.1% and 51.2%, respectively). Among children born outside an institution, 35.4% never had an HBR, compared to only 4.6% for institutional births.

Coverage of basic vaccines and drop-out ratios according to HBR ownership are presented in Figure 3. Analyses are restricted to children aged 12-23 months. These results must be interpreted with caution for the reasons laid out below in the Discussion. We found similar patterns for BCG and DPT1, as well as for DPT3 and Polio3, particularly for the two extreme categories - HBR seen and never had an HBR.

**FIGURE 3.**
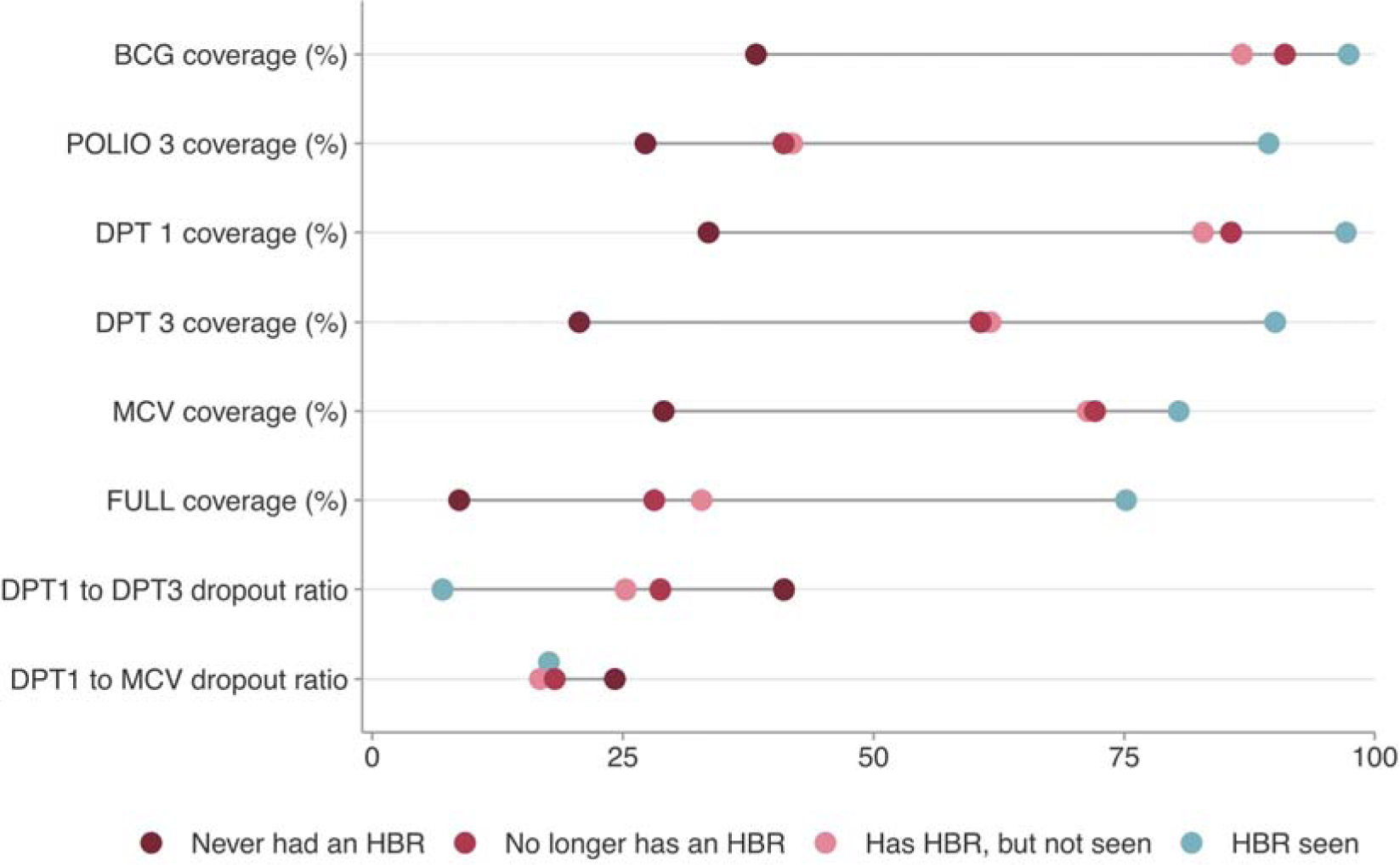
Coverage and drop-out of basic vaccines among children aged 12-23 months by HBR ownership.

When an HBR was seen, coverage of DPT1 and BCG were 97.2% and 97.5%, respectively. Among children who never had an HBR, coverage levels by recall were 28.7% and 33.0%, respectively. Similarly, DPT3 and Polio3 coverage were 90.5% and 89.7% among children with HBR seen, and 15.0% and 22.2% among those who never had an HBR.

MCV coverage among children with an HBR seen (80.3%) was lower than that for any other vaccines, including DPT3 and Polio3, suggesting drop-out between the third doses and MCV which is usually administered later. Results on dropout ratios are shown below. For children who never had an HBR, MCV coverage estimated by recall was 26.3%, similar to that for BCG and DPT1. For children who ever had an HBR (a combination of three categories), coverage was 95.5% for BCG, 94.2% for DPT1, 83.8% for DPT3, 79.1% for Ppolio3 and 78.7% for MCV.

Results for full vaccination reflect what was observed for each vaccine separately. Children for whom an HBR was seen had much higher coverage (75.5%) than those who never had an HBR (6.1%), whereas a smaller coverage gap was observed for those who no longer had an HBR (27.0%) compared to those who reportedly had an HBR which was not seen (37.4%). Among children who did not receive DPT1 (often referred to as zero-dose children), 71.3% (95% CI 69.8%-72.8%) had never had an HBR.

The dropout ratio between the first and third dose of DPT for children who never had an HBR was 47.6%; this information was based on recall of vaccinations. Among children whose HBR was seen, the dropout ratio was only 6.9%. For children who no longer had an HBR and those who reportedly had an HBR that was not seen, dropout ratios were 31.9% and 24.7%, respectively. We also investigated the proportions of children who failed to receive MCV among those who had received DPT1. Dropout was 24.5% for children who never had an HBR but was lower – ranging from 15.1% to 17.9% - in the other three HBR categories. Supplementary Table 3 provides the full results alongside their corresponding 95% confidence intervals. Supplementary Figure 1 provides the results for each of the 82 countries included.

## Discussion

We report on HBR ownership for children from 82 countries using data from national surveys carried out since 2010. Prior to our analyses, the largest multicountry publication on HBR ownership included 67 countries with surveys from 1993 to 2013. (9) We also report on within-country inequalities in HBR ownership by pooling results across all countries.

We found that many children did not have HBRs seen, but also that a significant proportion reported never having received one. The weighted prevalence of HBRs seen by the survey interviewer was 67.8%, whereas 10.2% had never received an HBR, and the remaining children had received one that was either lost or unavailable. We also found substantial variability by country and world regions, with median prevalence of HBRs seen ranging from about 90% in LMICs from Europe to under 60% in the Western Pacific region. Our results are consistent with those reported for 67 LMICs up to 2013, according to which HBRs were seen for over 80% of children in 23 countries and fewer than 50% in 24 countries.(9) According to the Global Health Observatory, current prevalence of HBRs seen across all countries with data ranged from 13% in Somalia to 100% in Belarus, Belgium, DPR Korea, and Turkmenistan.(5) Regarding world regions, this prevalence was lowest in the Western Pacific region (median of 42%), followed by the African region (66%). The highest prevalence was found in the European region (90%).

The availability of HBRs for inspection is dependent on several prerequisites.(18) First, HBRs must be available at national level, and then in the health facility. Next, the caregiver must receive it from a health worker usually during or before the first immunization encounter and bringing the record back to the facility when every new vaccine is administered. The record must be properly stored and looked after while in the household. For measurement purposes, during the household survey, the interviewer must request and wait to inspect the HBR to transcribe it. The home-based record must also be readable, and the interviewer must correctly transcribe vaccine dates or vaccination check marks. The increasing complexity of immunization schedules, the diverse range of home-based records (HBRs) in circulation, and the varying quality of their completion are making it increasingly challenging to record, read, and transcribe information from HBRs.(28–30)

Stockouts of HBRs have been documented to occur in many countries.(18). These stockouts can be attributed to various factors, including frequent changes to immunization schedules necessitating updates to HBRs, unclear responsibilities regarding printing and distribution, and rising costs associated with more advanced HBR designs, among other reasons.

In addition to stockouts, proper storage of the HBRs at home is an important issue. We found that for 9.2% of the children the HBR had been received but later lost, and for a larger proportion (12.8%) the HBR was reportedly still with the family but could not be produced. The loss of HBRs has been well documented;(31) this limits their potential to serve as a tool for continuity of care, particularly when individuals visit multiple health facilities. Lost and misplaced HBRs may preclude children from being immunized during visits to health centres, as suggested by studies from Ethiopia and Bangladesh.(32)

A framework was developed to guide the design and implementation of HBRs, as well as their distribution and the engagement of caregivers and users.(33) In particular, one cannot assume that failure to see an HBR in a survey solely reflects practices related to the caregiver. A survey from Kinshasa reported that lower proportions of HBRs were seen as field work advanced although the proportion of caregivers reporting ownership was maintained, which may be attributed to interviewer fatigue and inconsistent supervision.(34) Similarly, when comparing multiple-indicator health surveys with immunization-specific ones in close time proximity, the latter tend to report higher proportions of HBRs seen.(35) Unfortunately, the survey questionnaires and paradata used in the present analyses do not allow a more detailed examination of the reasons for reportedly having an HBR that could not be presented during the interview.

Analyses of inequalities help identify children without HBRs, thus contributing to public policies. Although children of different ages within the 6-35 months range have received HBRs to similar extents, the percentages of HBRs seen declined 0.7% per month of age. Sex of the child was not associated with ownership. Rural children were more likely than urban children to have ever had an HBR, but more likely to have lost or misplaced it, resulting in similar proportions of HBRs seen in both groups. As is the case for disparities in vaccine coverage,(12, 14) household wealth and maternal education are strongly associated with HBR prevalence; these differentials are particularly driven by never having received an HBR.

Just as immunization levels are strongly related to coverage of other essential primary health care interventions,(36) we found that HBR prevalence overlaps with antenatal and delivery care – particularly with the latter. The likely explanation is that some vaccines – most often BCG – are applied shortly after birth to children delivered in institutions. Over 95% of such children have ever received an HBR, compared to 74.6% of those born at home. Even though almost all children born in a facility received an HBR, 4.6% of them still failed to receive one. Stock-outs may be a potential reason for missing these children.(18) On the basis of the overlap between immunization coverage with indicators of multiple deprivation in terms of education, nutrition and sanitation(37), it is also likely that lack of HBRs will be frequent in such populations.

As expected, coverage with basic vaccines was highest among children whose HBRs were seen by surveyors and lowest among those who never had an HBR, a finding that has been previously reported.(35) Vaccine coverage - as ascertained through caregiver recall in children with lost or misplaced HBRs - was in between the two extreme categories of HBRs seen and never having had an HBR. Dropout ratios from DPT1 to DPT3 were as high as 50% for children who never received an HBR, compared to less than 10% among those whose HBRs were seen. DPT1-MCV dropout rates were more similar among the four HBR categories, ranging from 15.1% in the misplaced group and 24.5% among children who never had an HBR. These findings should not be interpreted as providing unidirectional evidence that HBR ownership will help increase coverage. Given that HBRs are provided at the time of the first vaccination – usually BCG shortly after birth – receiving an HBR and being vaccinated occur simultaneously. Low reported vaccine coverage among children who never received an HBR may reflect health systems failure in delivering immunization or alternatively caregivers forgetting past vaccinated received by their child. A review of the agreement between recall and documented vaccination showed that it varied significantly among settings and types of vaccines, with vaccines requiring multiple visits being the least reliable.(38) In settings with low HBR coverage, survey estimates of immunization rates should be regarded with caution.(8, 35)

Our analyses have limitations. Although over 80% of all low-income countries were included, standardized surveys were available for less than half of all upper-middle income countries. Eleven countries in which immunization HBRs are kept in health facilities were excluded from the analyses. Also, we are not able to explore HBR quality and accuracy of transcription when an HBR was seen. Another limitation is the lack of precision in separating reportedly lost HBRs from HBRs that were misplaced – that is, those that were still in possession of the family but were not seen by the surveyors. Both categories show similar associations with vaccine coverage and with sociodemographic and health services characteristics, which may be due to the fact that the two categories are indeed similar, or alternatively that surveys are unable to discriminate among misplaced and lost HBRs. A final limitation is that recall bias most likely affects data on coverage for children whose HBRs were not inspected.

## Conclusions

HBRs are important tools for several reasons.(1) They serve as a documentation tool that facilitates continuity of care and allows parents and health services to track the immunization status of a child, and to support compliance with vaccination schedules. HBRs allow healthcare providers to quickly determine which vaccines a child has already received and when they are due for the next vaccine, which has become even more important now as the IA2030 calls for a life-course approach to vaccination.(39) HBRs are also important for monitoring immunization coverage thought household surveys.

In the future, digitization of personal health records(40) may minimize the proportions of lost and misplaced HBRs, but such strategies will only be successful where internet access is equitable and widespread.

Our analyses identified countries and population subgroups for which low HBR coverage is likely leading to inaccurate and possibly biased estimates of immunization rates, and worse, likely affecting the likelihood of completing vaccination schedules. Regular monitoring of HBR ownership is essential for tracking trends and detecting subgroups of children needing special attention.

## Declarations

### Ethical aspects

Ethical clearance was the responsibility of the institutions that administered the surveys and all analyses relied on anonymized databases.

### Author contributions

All authors conceptualized the paper. BCP, AW, TMS and LA conducted the analyses and verified the underlying data, with support from CGV and AJDB. All authors interpreted the results. MCDH, CGV and BCP prepared the first draft of the manuscript, which was revised and edited by all other authors. All authors read and approved the final manuscript. All authors attest they meet the ICMJE criteria for authorship.

## Acknowledgements

This paper was made possible with funds from Bill & Melinda Gates Foundation (grant number OPP1199234), Gavi, the Vaccine Alliance (grant number N/A) and Associação Brasileira de Saúde Coletiva (grant number N/A).

## Data availability Statement

All the analyses were carried out using publicly available datasets that can be obtained directly from the DHS (dhsprogram.com) and the MICS (mics.unicef.org) websites.

