## Supplementary table for "Inequalities in ownership and availability of home-based vaccination records in 82 low- and middle-income countries"

### Supplementary materials

ST 1 – Surveys included in the analyses.

| ISO code | Country | Year | Source |
|---|---|---|---|
| AFG | Afghanistan | 2015 | DHS |
| DZA | Algeria | 2018 | MICS |
| AGO | Angola | 2015 | DHS |
| ARM | Armenia | 2015 | DHS |
| BGD | Bangladesh | 2017 | DHS |
| BLZ | Belize | 2015 | MICS |
| BEN | Benin | 2017 | DHS |
| BIH | Bosnia and Herzegovina | 2011 | MICS |
| BFA | Burkina Faso | 2010 | DHS |
| BDI | Burundi | 2016 | DHS |
| CAF | CAR | 2018 | MICS |
| KHM | Cambodia | 2014 | DHS |
| CMR | Cameroon | 2018 | DHS |
| TCD | Chad | 2014 | DHS |
| COL | Colombia | 2010 | DHS |
| COM | Comoros | 2012 | DHS |
| COG | Congo Brazzaville | 2014 | MICS |
| COD | Congo Democratic Republic | 2017 | MICS |
| CRI | Costa Rica | 2018 | MICS |
| CIV | Cote dIvoire | 2016 | MICS |
| DOM | Dominican Republic | 2019 | MICS |
| EGY | Egypt | 2014 | DHS |
| SLV | El Salvador | 2014 | MICS |
| SWZ | Eswatini | 2014 | MICS |
| FJI | Fiji | 2021 | MICS |
| GAB | Gabon | 2012 | DHS |
| GMB | Gambia | 2019 | DHS |
| GHA | Ghana | 2017 | MICS |
| GTM | Guatemala | 2014 | DHS |
| GIN | Guinea | 2018 | DHS |
| GNB | Guinea Bissau | 2018 | MICS |
| GUY | Guyana | 2019 | MICS |
| HTI | Haiti | 2016 | DHS |
| HND | Honduras | 2019 | MICS |
| IND | India | 2019 | DHS |
| IDN | Indonesia | 2017 | DHS |
| IRQ | Iraq | 2018 | MICS |
| JAM | Jamaica | 2011 | MICS |
| JOR | Jordan | 2017 | DHS |
| KEN | Kenya | 2014 | DHS |
| KIR | Kiribati | 2018 | MICS |
| XKX | Kosovo | 2019 | MICS |
| LAO | Lao | 2017 | MICS |

| | | | |
|---|---|---|---|
| LSO | Lesotho | 2018 | MICS |
| LBR | Liberia | 2019 | DHS |
| MDG | Madagascar | 2021 | DHS |
| MWI | Malawi | 2019 | MICS |
| MDV | Maldives | 2016 | DHS |
| MLI | Mali | 2018 | DHS |
| MRT | Mauritania | 2019 | DHS |
| MEX | Mexico | 2015 | MICS |
| MNE | Montenegro | 2013 | MICS |
| MOZ | Mozambique | 2015 | DHS |
| MMR | Myanmar | 2015 | DHS |
| NAM | Namibia | 2013 | DHS |
| NPL | Nepal | 2019 | MICS |
| NER | Niger | 2021 | DHS |
| NGA | Nigeria | 2018 | DHS |
| PAK | Pakistan | 2017 | DHS |
| PNG | Papua New Guinea | 2016 | DHS |
| PRY | Paraguay | 2016 | MICS |
| PER | Peru | 2020 | DHS |
| PHL | Philippines | 2017 | DHS |
| RWA | Rwanda | 2019 | DHS |
| WSM | Samoa | 2019 | MICS |
| STP | Sao Tome and Principe | 2019 | MICS |
| SEN | Senegal | 2019 | DHS |
| SLE | Sierra Leone | 2019 | DHS |
| ZAF | South Africa | 2016 | DHS |
| PSE | State of Palestine | 2019 | MICS |
| TJK | Tajikistan | 2017 | DHS |
| TZA | Tanzania | 2015 | DHS |
| THA | Thailand | 2019 | MICS |
| TLS | Timor Leste | 2016 | DHS |
| TGO | Togo | 2017 | MICS |
| TUN | Tunisia | 2018 | MICS |
| TUR | Türkiye | 2013 | DHS |
| TUV | Tuvalu | 2019 | MICS |
| UGA | Uganda | 2016 | DHS |
| YEM | Yemen | 2013 | DHS |
| ZMB | Zambia | 2018 | DHS |
| ZWE | Zimbabwe | 2019 | MICS |

ST 2 – Percentages of children aged 6-35 months according to home-based record ownership status stratified by age groups, wealth quintiles, mother's education, area of residence, antenatal care and institutional delivery

| Group | Level | Card status | % Children | 95% CI | |
|---|---|---|---|---|---|
| All children | 6-35 months | Never had a card | 10.2% | 9.9% | 10.5% |
| All children | 6-35 months | Card seen | 67.8% | 67.4% | 68.2% |
| All children | 6-35 months | Has card, but not seen | 12.8% | 12.5% | 13.0% |
| All children | 6-35 months | No longer has a card | 9.2% | 9.0% | 9.4% |
| Age group | 6-11 months | Never had a card | 9.5% | 9.0% | 9.9% |
| Age group | 6-11 months | Card seen | 76.3% | 75.7% | 76.9% |
| Age group | 6-11 months | Has card, but not seen | 9.1% | 8.7% | 9.5% |
| Age group | 6-11 months | No longer has a card | 5.2% | 4.9% | 5.5% |
| Age group | 12-23 months | Never had a card | 9.7% | 9.3% | 10.1% |
| Age group | 12-23 months | Card seen | 69.9% | 69.4% | 70.4% |
| Age group | 12-23 months | Has card, but not seen | 11.9% | 11.6% | 12.2% |
| Age group | 12-23 months | No longer has a card | 8.5% | 8.2% | 8.8% |
| Age group | 24-35 months | Never had a card | 11.1% | 10.7% | 11.5% |
| Age group | 24-35 months | Card seen | 61.5% | 60.9% | 62.0% |
| Age group | 24-35 months | Has card, but not seen | 15.5% | 15.1% | 15.8% |
| Age group | 24-35 months | No longer has a card | 12.0% | 11.6% | 12.3% |
| Sex | Male | Never had a card | 10.0% | 9.6% | 10.3% |
| Sex | Female | Never had a card | 10.5% | 10.1% | 10.8% |
| Sex | Male | Card seen | 68.3% | 67.8% | 68.8% |
| Sex | Female | Card seen | 67.3% | 66.8% | 67.8% |
| Sex | Male | Has card, but not seen | 12.7% | 12.4% | 13.0% |
| Sex | Female | Has card, but not seen | 12.8% | 12.5% | 13.2% |
| Sex | Male | No longer has a card | 9.0% | 8.7% | 9.3% |
| Sex | Female | No longer has a card | 9.4% | 9.1% | 9.7% |
| Wealth quintiles | Poorest | Never had a card | 17.1% | 16.3% | 17.9% |
| Wealth quintiles | Second | Never had a card | 12.4% | 11.8% | 13.0% |
| Wealth quintiles | Third | Never had a card | 9.4% | 8.8% | 10.0% |
| Wealth quintiles | Fourth | Never had a card | 6.1% | 5.6% | 6.6% |
| Wealth quintiles | Wealthiest | Never had a card | 3.7% | 3.4% | 4.1% |
| Wealth quintiles | Poorest | Card seen | 62.4% | 61.6% | 63.1% |
| Wealth quintiles | Second | Card seen | 67.1% | 66.4% | 67.8% |
| Wealth quintiles | Third | Card seen | 69.4% | 68.7% | 70.1% |
| Wealth quintiles | Fourth | Card seen | 70.8% | 70.0% | 71.7% |
| Wealth quintiles | Wealthiest | Card seen | 70.8% | 70.0% | 71.6% |
| Wealth quintiles | Poorest | Has card, but not seen | 11.5% | 11.1% | 11.9% |
| Wealth quintiles | Second | Has card, but not seen | 11.6% | 11.2% | 12.1% |
| Wealth quintiles | Third | Has card, but not seen | 12.3% | 11.8% | 12.8% |
| Wealth quintiles | Fourth | Has card, but not seen | 13.6% | 13.0% | 14.2% |
| Wealth quintiles | Wealthiest | Has card, but not seen | 15.5% | 14.9% | 16.2% |
| Wealth quintiles | Poorest | No longer has a card | 9.1% | 8.6% | 9.5% |

| Group | Level | Card status | % Children | 95% CI | |
|---|---|---|---|---|---|
| Wealth quintiles | Second | No longer has a card | 8.9% | 8.5% | 9.3% |
| Wealth quintiles | Third | No longer has a card | 8.9% | 8.5% | 9.3% |
| Wealth quintiles | Fourth | No longer has a card | 9.5% | 9.0% | 10.0% |
| Wealth quintiles | Wealthiest | No longer has a card | 9.9% | 9.4% | 10.5% |
| Mother’s education | None | Never had a card | 23.0% | 22.1% | 24.0% |
| Mother’s education | Primary | Never had a card | 10.7% | 10.2% | 11.2% |
| Mother’s education | Secondary+ | Never had a card | 4.3% | 4.1% | 4.6% |
| Mother’s education | None | Card seen | 55.7% | 54.9% | 56.6% |
| Mother’s education | Primary | Card seen | 67.2% | 66.5% | 67.9% |
| Mother’s education | Secondary+ | Card seen | 73.4% | 72.9% | 73.9% |
| Mother’s education | None | Has card, but not seen | 9.5% | 9.1% | 9.9% |
| Mother’s education | Primary | Has card, but not seen | 12.1% | 11.7% | 12.5% |
| Mother’s education | Secondary+ | Has card, but not seen | 14.5% | 14.1% | 14.8% |
| Mother’s education | None | No longer has a card | 11.7% | 11.3% | 12.2% |
| Mother’s education | Primary | No longer has a card | 10.0% | 9.6% | 10.4% |
| Mother’s education | Secondary+ | No longer has a card | 7.8% | 7.5% | 8.1% |
| Area of residence | Urban | Never had a card | 6.3% | 5.9% | 6.7% |
| Area of residence | Rural | Never had a card | 12.4% | 12.0% | 12.9% |
| Area of residence | Urban | Card seen | 68.3% | 67.7% | 69.0% |
| Area of residence | Rural | Card seen | 67.5% | 67.0% | 68.0% |
| Area of residence | Urban | Has card, but not seen | 15.2% | 14.8% | 15.7% |
| Area of residence | Rural | Has card, but not seen | 11.3% | 11.1% | 11.6% |
| Area of residence | Urban | No longer has a card | 10.1% | 9.7% | 10.5% |
| Area of residence | Rural | No longer has a card | 8.7% | 8.4% | 9.0% |
| Antenatal care | 4+ visits | Never had a card | 4.7% | 4.5% | 5.0% |
| Antenatal care | <4 visits | Never had a card | 17.3% | 16.7% | 17.9% |
| Antenatal care | 4+ visits | Card seen | 73.2% | 72.8% | 73.6% |
| Antenatal care | <4 visits | Card seen | 63.1% | 62.5% | 63.8% |
| Antenatal care | 4+ visits | Has card, but not seen | 13.8% | 13.5% | 14.1% |
| Antenatal care | <4 visits | Has card, but not seen | 10.6% | 10.2% | 10.9% |
| Antenatal care | 4+ visits | No longer has a card | 8.2% | 8.0% | 8.5% |
| Antenatal care | <4 visits | No longer has a card | 9.0% | 8.7% | 9.4% |
| Institutional delivery | Institutional | Never had a card | 4.6% | 4.4% | 4.8% |
| Institutional delivery | Noninstitutional | Never had a card | 25.4% | 24.6% | 26.3% |
| Institutional delivery | Institutional | Card seen | 74.1% | 73.7% | 74.4% |
| Institutional delivery | Noninstitutional | Card seen | 51.2% | 50.4% | 52.0% |
| Institutional delivery | Institutional | Has card, but not seen | 13.2% | 12.9% | 13.4% |
| Institutional delivery | Noninstitutional | Has card, but not seen | 11.6% | 11.2% | 12.0% |
| Institutional delivery | Institutional | No longer has a card | 8.2% | 7.9% | 8.4% |
| Institutional delivery | Noninstitutional | No longer has a card | 11.8% | 11.4% | 12.3% |
| Age group | 6-11 months | Never had a card | 12.9% | 9.8% | 16.8% |
| Age group | 6-11 months | Card seen | 83.2% | 79.5% | 86.3% |
| Age group | 6-11 months | Has card, but not seen | 1.5% | 1.0% | 2.1% |
| Age group | 6-11 months | No longer has a card | 2.4% | 1.3% | 4.5% |

| Group | Level | Card status | % Children | 95% CI | |
|---|---|---|---|---|---|
| Age group | 12-23 months | Never had a card | 12.1% | 9.4% | 15.4% |
| Age group | 12-23 months | Card seen | 83.8% | 80.0% | 87.1% |
| Age group | 12-23 months | Has card, but not seen | 2.0% | 1.4% | 2.8% |
| Age group | 12-23 months | No longer has a card | 2.1% | 1.2% | 3.4% |
| Age group | 24-35 months | Never had a card | 15.7% | 12.6% | 19.5% |
| Age group | 24-35 months | Card seen | 79.8% | 75.6% | 83.4% |
| Age group | 24-35 months | Has card, but not seen | 2.1% | 1.6% | 2.8% |
| Age group | 24-35 months | No longer has a card | 2.4% | 1.5% | 3.7% |
| All children | 6-35 months | Never had a card | 13.7% | 11.5% | 16.3% |
| All children | 6-35 months | Card seen | 82.1% | 79.2% | 84.6% |
| All children | 6-35 months | Has card, but not seen | 1.9% | 1.6% | 2.4% |
| All children | 6-35 months | No longer has a card | 2.3% | 1.6% | 3.2% |
| Sex | Male | Never had a card | 12.6% | 10.0% | 15.7% |
| Sex | Female | Never had a card | 14.9% | 12.3% | 18.0% |
| Sex | Male | Card seen | 83.5% | 80.2% | 86.3% |
| Sex | Female | Card seen | 80.6% | 77.1% | 83.7% |
| Sex | Male | Has card, but not seen | 1.9% | 1.5% | 2.5% |
| Sex | Female | Has card, but not seen | 1.9% | 1.4% | 2.6% |
| Sex | Male | No longer has a card | 2.0% | 1.2% | 3.3% |
| Sex | Female | No longer has a card | 2.5% | 1.7% | 3.7% |
| Wealth quintiles | Poorest | Never had a card | 25.4% | 21.2% | 30.2% |
| Wealth quintiles | Second | Never had a card | 14.9% | 10.6% | 20.5% |
| Wealth quintiles | Third | Never had a card | 11.2% | 8.0% | 15.4% |
| Wealth quintiles | Fourth | Never had a card | 9.4% | 5.9% | 14.5% |
| Wealth quintiles | Wealthiest | Never had a card | 6.9% | 4.1% | 11.5% |
| Wealth quintiles | Poorest | Card seen | 71.0% | 65.6% | 75.8% |
| Wealth quintiles | Second | Card seen | 81.9% | 76.6% | 86.2% |
| Wealth quintiles | Third | Card seen | 85.1% | 80.6% | 88.8% |
| Wealth quintiles | Fourth | Card seen | 85.8% | 81.2% | 89.5% |
| Wealth quintiles | Wealthiest | Card seen | 87.3% | 81.6% | 91.5% |
| Wealth quintiles | Poorest | Has card, but not seen | 1.9% | 1.2% | 3.0% |
| Wealth quintiles | Second | Has card, but not seen | 1.6% | 1.0% | 2.5% |
| Wealth quintiles | Third | Has card, but not seen | 1.7% | 1.0% | 2.8% |
| Wealth quintiles | Fourth | Has card, but not seen | 1.8% | 1.2% | 2.6% |
| Wealth quintiles | Wealthiest | Has card, but not seen | 2.7% | 1.9% | 3.6% |
| Wealth quintiles | Poorest | No longer has a card | 1.7% | 0.9% | 3.2% |
| Wealth quintiles | Second | No longer has a card | 1.6% | 0.9% | 3.0% |
| Wealth quintiles | Third | No longer has a card | 2.0% | 1.1% | 3.7% |
| Wealth quintiles | Fourth | No longer has a card | 3.0% | 1.6% | 5.4% |
| Wealth quintiles | Wealthiest | No longer has a card | 3.1% | 1.8% | 5.3% |
| Mother’s education | None | Never had a card | 33.7% | 28.2% | 39.7% |
| Mother’s education | Primary | Never had a card | 17.3% | 13.2% | 22.4% |
| Mother’s education | Secondary+ | Never had a card | 1.6% | 1.0% | 2.6% |
| Mother’s education | None | Card seen | 62.4% | 56.6% | 67.9% |

| Group | Level | Card status | % Children | 95% CI | |
|---|---|---|---|---|---|
| Mother’s education | Primary | Card seen | 77.6% | 71.7% | 82.6% |
| Mother’s education | Secondary+ | Card seen | 94.4% | 93.3% | 95.3% |
| Mother’s education | None | Has card, but not seen | 0.5% | 0.2% | 1.1% |
| Mother’s education | Primary | Has card, but not seen | 0.8% | 0.3% | 2.2% |
| Mother’s education | Secondary+ | Has card, but not seen | 3.2% | 2.6% | 3.8% |
| Mother’s education | None | No longer has a card | 3.4% | 1.8% | 6.3% |
| Mother’s education | Primary | No longer has a card | 4.2% | 2.6% | 6.7% |
| Mother’s education | Secondary+ | No longer has a card | 0.8% | 0.4% | 1.6% |
| Area of residence | Urban | Never had a card | 8.7% | 5.5% | 13.3% |
| Area of residence | Rural | Never had a card | 16.3% | 13.4% | 19.7% |
| Area of residence | Urban | Card seen | 85.4% | 80.2% | 89.4% |
| Area of residence | Rural | Card seen | 80.4% | 76.8% | 83.6% |
| Area of residence | Urban | Has card, but not seen | 3.2% | 2.6% | 4.0% |
| Area of residence | Rural | Has card, but not seen | 1.3% | 0.9% | 1.8% |
| Area of residence | Urban | No longer has a card | 2.8% | 1.6% | 4.9% |
| Area of residence | Rural | No longer has a card | 2.0% | 1.3% | 3.0% |
| Antenatal care | 4+ visits | Never had a card | 4.1% | 2.8% | 5.9% |
| Antenatal care | <4 visits | Never had a card | 32.1% | 27.2% | 37.4% |
| Antenatal care | 4+ visits | Card seen | 91.4% | 88.9% | 93.4% |
| Antenatal care | <4 visits | Card seen | 64.5% | 59.1% | 69.5% |
| Antenatal care | 4+ visits | Has card, but not seen | 2.1% | 1.5% | 2.8% |
| Antenatal care | <4 visits | Has card, but not seen | 0.7% | 0.3% | 1.4% |
| Antenatal care | 4+ visits | No longer has a card | 2.4% | 1.5% | 4.0% |
| Antenatal care | <4 visits | No longer has a card | 2.8% | 1.7% | 4.6% |
| Institutional delivery | Institutional | Never had a card | 5.8% | 4.2% | 7.9% |
| Institutional delivery | Noninstitutional | Never had a card | 39.2% | 33.8% | 44.9% |
| Institutional delivery | Institutional | Card seen | 90.4% | 88.0% | 92.3% |
| Institutional delivery | Noninstitutional | Card seen | 56.0% | 49.7% | 62.2% |
| Institutional delivery | Institutional | Has card, but not seen | 1.8% | 1.4% | 2.3% |
| Institutional delivery | Noninstitutional | Has card, but not seen | 0.9% | 0.4% | 2.1% |
| Institutional delivery | Institutional | No longer has a card | 2.1% | 1.3% | 3.4% |
| Institutional delivery | Noninstitutional | No longer has a card | 3.9% | 2.5% | 6.0% |

ST 3 – Immunization coverage among children aged 12-23 months according to home-based record ownership status.

| Vaccine | HBR status | Coverage | 95% CI | |
|---|---|---|---|---|
| BCG | Never had a card | 33.0% | 31.5% | 34.6% |
| BCG | Card seen | 97.5% | 97.4% | 97.7% |
| BCG | Has card, but not seen | 87.6% | 86.6% | 88.5% |
| BCG | No longer has a card | 89.8% | 88.9% | 90.6% |
| POLIO 3 | Never had a card | 22.2% | 20.7% | 23.9% |
| POLIO 3 | Card seen | 89.7% | 89.4% | 90.0% |
| POLIO 3 | Has card, but not seen | 45.8% | 44.4% | 47.2% |
| POLIO 3 | No longer has a card | 39.9% | 38.0% | 41.9% |
| DPT 3 | Never had a card | 15.0% | 13.9% | 16.2% |
| DPT 3 | Card seen | 90.5% | 90.2% | 90.8% |
| DPT 3 | Has card, but not seen | 63.4% | 62.0% | 64.7% |
| DPT 3 | No longer has a card | 57.6% | 55.8% | 59.4% |
| DPT 1 | Never had a card | 28.7% | 27.2% | 30.2% |
| DPT 1 | Card seen | 97.2% | 97.0% | 97.3% |
| DPT 1 | Has card, but not seen | 84.1% | 83.1% | 85.0% |
| DPT 1 | No longer has a card | 84.5% | 83.3% | 85.6% |
| MCV | Never had a card | 26.3% | 24.9% | 27.7% |
| MCV | Card seen | 80.3% | 79.8% | 80.7% |
| MCV | Has card, but not seen | 73.6% | 72.4% | 74.8% |
| MCV | No longer has a card | 72.7% | 71.1% | 74.1% |
| FULL | Never had a card | 6.1% | 5.5% | 6.8% |
| FULL | Card seen | 75.5% | 75.1% | 75.9% |
| FULL | Has card, but not seen | 37.4% | 36.1% | 38.8% |
| FULL | No longer has a card | 27.0% | 25.3% | 28.7% |
| DPT dropout ratio | Never had a card | 47.6% | 44.7% | 50.6% |
| DPT dropout ratio | Card seen | 6.9% | 6.6% | 7.1% |
| DPT dropout ratio | Has card, but not seen | 24.7% | 23.3% | 26.0% |
| DPT dropout ratio | No longer has a card | 31.9% | 30.0% | 33.8% |

Legend: HBR – home-based record; BCG – Bacille Calmette-Guérin; DPT – diphtheria-pertussis-tetanus; MCV – measles containing vaccine; FULL – full immunization coverage (BCG + DPT3 + Polio3 + MCV)

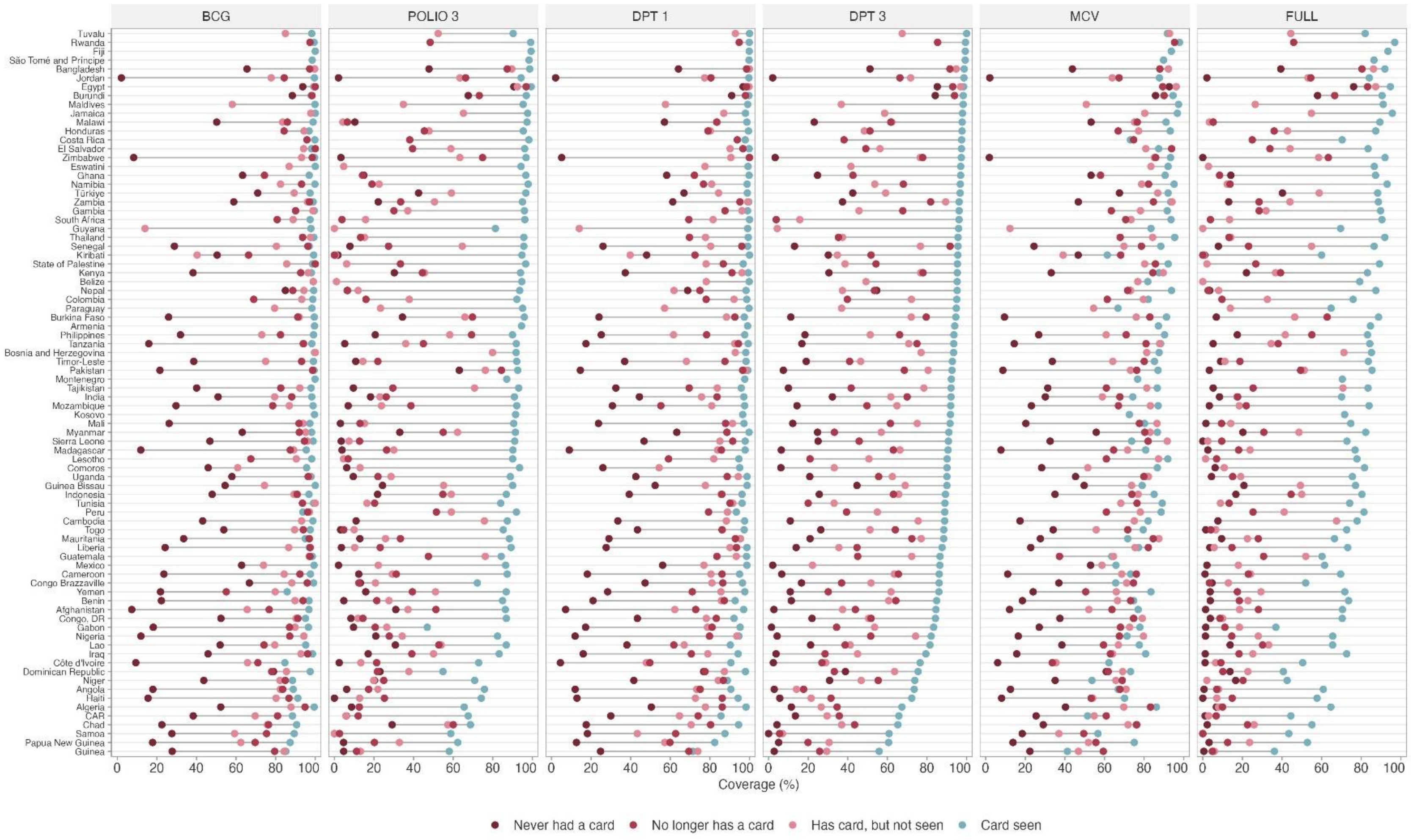


SF 1 – Immunization coverage among children aged 12-23 months according to home-based record ownership status by country. Countries ordered according to DPT3 coverage.
